# Development of an automated chemiluminescence assay system for quantitative measurement of multiple anti-SARS-CoV-2 antibodies

**DOI:** 10.1101/2020.11.04.20225805

**Authors:** Sousuke Kubo, Norihisa Ohtake, Kei Miyakawa, Sundararaj Stanleyraj Jeremiah, Yutaro Yamaoka, Kota Murohashi, Eri Hagiwara, Takahiro Mihara, Atsushi Goto, Etsuko Yamazaki, Takashi Ogura, Takeshi Kaneko, Takeharu Yamanaka, Akihide Ryo

**Affiliations:** Department of Microbiology, Yokohama City University Graduate School of Medicine, Kanagawa, Japan; Department of Pulmonology, Yokohama City University Graduate School of Medicine, Kanagawa, Japan; Bioscience Division, Reagent Development Department, Tosoh Corporation, Kanagawa, Japan; Advanced Medical Research Center, Yokohama City University, Kanagawa, Japan; Life Science Laboratory, Technology and Development Division, Kanto Chemical Co, Inc, Kanagawa, Japan; Department of Respiratory Medicine, Kanagawa Cardiovascular and Respiratory Center, Kanagawa, Japan; Department of Health Data Science, Yokohama City University Graduate School of Data Science, Kanagawa, Japan; Clinical Laboratory Department, Yokohama City University Hospital, Kanagawa, Japan; Department of Biostatistics, Yokohama City University Graduate School of Medicine, Kanagawa, Japan

## Abstract

**Objective:** Serological tests for COVID-19 have been instrumental in studying the epidemiology of the disease. However, the performance of the currently available tests is plagued by the problem of variability. We have developed a high-throughput serological test capable of simultaneously detecting total immunoglobulins (Ig) and immunoglobulin G (IgG) against two of the most immunologically relevant SARS-CoV-2 antigens, nucleocapsid protein (NP) and spike protein (SP) and report its performance in detecting COVID-19 in clinical samples.

**Methods:** We designed and prepared reagents for measuring NP-IgG, NP-Total Ig, SP-IgG, and SP-Total Ig (using N-terminally truncated NP (ΔN-NP) or receptor-binding domain (RBD) antigen) on the advanced chemiluminescence enzyme immunoassay system TOSOH AIA-CL. After determining the basal thresholds based on 17 sera obtained from confirmed COVID-19 patients and 600 negative sera. Subsequently, the clinical validity of the assay was evaluated using independent 202 positive samples and 1,000 negative samples from healthy donors.

**Results:** All of the four test parameters showed 100% specificity individually (1,000/1,000; 95%CI, 99.63-100). The sensitivity of the assay increased proportionally to the elapsed time from symptoms onset, and all the tests achieved 100% sensitivity (153/153; 95%CI, 97.63-100) after 13 days from symptoms onset. NP-Total Ig was the earliest to attain maximal sensitivity among the other antibodies tested.

**Conclusion:** Our newly developed serological testing exhibited 100% sensitivity and specificity after 13 days from symptoms onset. Hence, it could be used as a reliable method for accurate detection of COVID-19 patients and to evaluate seroprevalence and possibly for surrogate assessment of herd immunity.

## Introduction

Severe acute respiratory syndrome coronavirus 2 (SARS-CoV-2) is the causative agent of the coronavirus disease 2019 (COVID-19). The ongoing global pandemic caused by SARS-CoV-2 is one of the most serious crises that humanity has faced in the recent past. COVID-19 infections require intensive treatment in 20–25% of patients and result in death of 4–11% of hospitalized patients (1). Serological tests are particularly helpful in the detection of ongoing COVID-19 infection, depending on the time course of the illness. Moreover, serological assays enable to estimate the prevalence of existing immunity to SARS-CoV-2 in a population (2).

In general, antibody testing not only helps to indirectly diagnose an infection but also reflects the individual immune response (3). This is fundamentally different from reverse transcription polymerase chain reaction (RT-PCR) or antigen detection tests, which measure viral nucleic acids and antigens, respectively. Unlike other infections, COVID-19 exhibits concurrent elevation of immunoglobulin (Ig) G and IgM in the second week after disease onset, with IgM reaching peak titers in the second week and IgG in the third week of the disease (4, 5). Since the transition of antibody titers over time is different in different types of immunoglobulin, it is recognized that the measurement of multiple antibody titers can holistically assess the immune status in infected individuals. Various antigens of SARS-CoV-2 can be utilized by serodiagnostic kits for the detection of virus-specific antibodies. These include viral surface antigens such as membrane proteins, envelope proteins, and spike protein (SP) and non-surface antigens such as the nucleocapsid protein (NP) (6, 7). Of these, the SP and the NP are the most widely employed for serological diagnosis (8). The SP mediates viral entry into host cells through the receptor-binding domain (RBD) and antibodies against RBD are predicted to represent the protective immunity to SARS-CoV-2 infection (9, 10). The NP not only participates in ribonucleic acid (RNA) packaging and virus particle release but also plays a principal role in orchestrating various intracellular events required for viral replication after entry (11). The NP shares antigenic similarity with other human coronaviruses and may exhibit cross-reactivity when used in serological tests as the full-length protein, but is highly specific for SARS-CoV-2 in the absence of its N-terminal amino acid residues (12).

High-throughput COVID-19 serology testing is required for epidemiological surveillance. The most common method for measuring antibody titers is enzyme-linked immunosorbent assay (ELISA). Although ELISA has many advantages, it is a time-consuming and labor-intensive procedure with a higher likelihood of technical errors than other automated platforms (13). To overcome these problems, we used the automated chemiluminescent enzyme immunoassay system AIA-CL (TOSOH, Tokyo, Japan), to develop a serodiagnostic test that quantitatively detects SARS-CoV-2 antibody titers with a short turn-around-time of 10 minutes. Apart from lower manual errors, this platform also has the inherent advantage of being more precise and sensitive than ELISA (14). We evaluated the performance of the AIA-CL, and analyzed chronological change of antibody titers in patients.

## Methods

### Antigen design

Previously, we showed that the cross-reactivity of NP is due to conserved residues in its N-terminus; accordingly, truncated N-terminal NP (ΔN-NP) is highly specific for detection of SARS-CoV-2. ΔN-NP (121–419 aa) was synthesized using the wheat germ cell–free protein synthesis system and purified using Ni-Sepharose beads (15). The complementary deoxyribonucleic acid (cDNA) sequence corresponding to the RBD (residues 319-541) of SARS-CoV-2 spike protein was chemically synthesized and inserted into in-house Strep-tagged mammalian expression vector. Strep-tagged RBD protein was transient expressed in a mammalian cell (Expi293, Thermo Fisher Scientific), purified by Strep-tag purification system (IBA) (16).

### Development of an antibody detection reagent for AIA-CL analyzers

We developed four types of serological test reagents (TOSOH NP-IgG, NP-Total Ig, SP-IgG, SP-Total Ig test) to detect anti–SARS-CoV-2 antibodies on the AIA-CL. The serological test comprises of an automated immunoassay for the quantitative determination of either serum IgG or Total Ig (IgG, IgM, IgA, IgE, etc.) levels against the ΔN-NP or RBD antigen. Micro-magnetic beads coated with ΔN-NP or RBD antigen were added to the first reaction layer, and alkaline phosphatase–conjugated anti-human IgG or ΔN-NP antigen or RBD antigen was added to the second reaction layer and prepared by an immediate lyophilization in a test cup. The AIA-CL system performs an automated process that includes specimen dispensing, incubation of the reaction cup, bound/free washing procedure, dispensing of in-house chemiluminescent substrates (DIFURAT), chemiluminescence detection, and reporting of results (up to 240 tests/hour/device).

### Patient cohorts

For initial performance evaluation, 600 samples of healthy donors collected prior to the spread of COVID-19 infection (purchased from TRINA BIOREACTIVES AG, Zurich, Swiss) and 17 samples from COVID-19 antibody–positive patients (according to the Abbott Architect SARS-CoV-2 IgG antibody test) were purchased from Biomex GmbH (Heidelberg, German). To examine the correlation between the results of the NP-IgG and NP-Total Ig tests from this study with those of other commercially available kits, 44 samples from COVID-19 antibody–positive patients were purchased from Biomex GmbH. Since there was no quantitative kit of antibodies against SP available from other companies, we did not compare the result of SP-IgG and SP-Total Ig with other companies. For calculation of clinical sensitivity and specificity, a total of 202 samples from 42 symptomatic patients with PCR-confirmed SARS-CoV-2 infection hospitalized in Yokohama City University Hospital (YCUH) and Kanagawa Cardiovascular and Respiratory Center (KCRC) were used. The severity of symptoms on admission was determined according to National Institutes of Health (NIH) classification (17). If the patient was in the recovery phase (more than 30 days after symptoms onset and the symptoms were improving on admission), categorized as “others”. To ensure the quality of the clinical specimens, one patient with a history of anticancer therapy was excluded. One thousand samples of healthy donors collected prior to the spread of COVID-19 infection (provided by the Biobank Division of Yokohama City University Advanced Medical Research Center) were used as negative controls.

Mann-Whitney Rank Sum test was performed to identify the correlations of health donors and COVID-19 patients. In the initial evaluation, we set the cutoff values of NP-IgG, NP-Total Ig, SP-IgG and SP-Total Ig titers by maximizing the overall prediction performance with the Youden’s J index (J = sensitivity + specificity **−** 1) based on the receiver operating characteristic (ROC) analysis, and defined them as 1.0 index. Next, the sensitivity, specificity and 95% confidence interval (95% CI) of the clinical samples was calculated using ROC curve. A p-value of <0.05 was considered to be indicative of statistical significance. All statistical analyses were performed by using GraphPad Prism 6 software.

All healthy donors provided written informed consent for use of their clinical and biological data for the purpose of the scientific research. All patients were recruited using the opt-out approaches. The study was performed according to protocols approved by our institutional review board.

## Results

### Preparation of reagents and determination of the optimal cutoff value

Reagents for the NP-IgG, NP-Total Ig, SP-IgG and SP-Total Ig tests were prepared using ΔN-NP and RBD antigen respectively. Schematic illustration of the assay principles used in the TOSOH AIA-CL system is depicted in Figure 1. After confirming within-run reproducibility, we measured the intensity of the chemiluminescent signals of 600 healthy donor samples and 17 COVID-19 antibody–positive samples (Figure S1 A-D). Using the ROC curve, we determined the cutoff values at 3,889 counts of intensity for NP-IgG, 1,705 counts for NP-Total Ig, 6,700 counts for SP-IgG, and 1,000 counts for SP-Total Ig, and defined them as 1.0 index (Figure S1 E-H).

**Figure 1.**
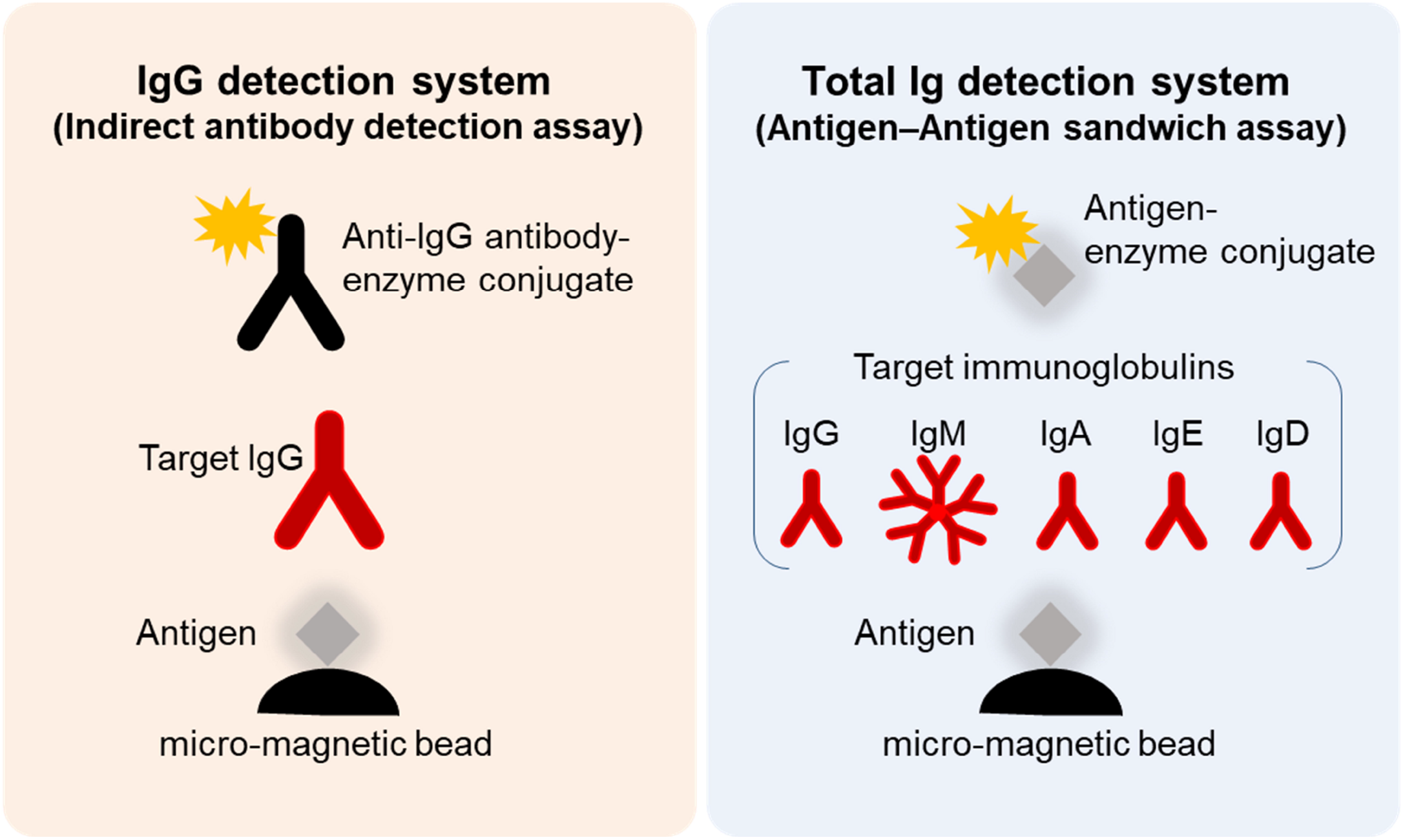
Schematic illustration of SARS-CoV-2 NP-IgG, NP-total Ig, SP-IgG and SP-Total Ig tests in the TOSOH AIA-CL system. Measurement method of the NP-IgG, NP-Total Ig, SP-IgG and SP-Total Ig tests in the AIA-CL system. Micro-magnetic beads coated with ΔN-NP or RBD antigen were added to the first reaction layer. NP-IgG or SP-IgG in patient’s serum was captured by alkaline phosphatase–conjugated anti–human IgG secondary antibody (left panel). NP-Total Ig or SP-Total Ig was captured by alkaline phosphatase–conjugated ΔN-NP or RBD antigen (right panel). Alkaline phosphatase, used as the labeling enzyme, was detected using in-house chemiluminescent substrates (DIFURAT). NP: nucleocapsid protein, SP: spike protein, AIA-CL: automated chemiluminescent enzyme immunoassay system, RBD: receptor-binding domain.

### Chronological analysis of antibody titers

Next, we evaluated a total of 202 samples collected after 7 days from symptoms onset from 43 symptomatic patients with COVID-19 diagnosed by RT-PCR, using the NP-IgG, NP-Total Ig, SP-IgG, SP-Total Ig reagents. The median age of COVID-19 patients was 61.3 years in YCUH and 64.5 years in KCRC. 2 patients were mild cases, 17 were moderate cases, 16 were severe cases, 7 were critical cases, and 2 were categorized as “others” on admission (Table S1). We compared antibody titers in the following cohorts: 7–9 days, 10–12 days, 13–20 days, 21–30 days, and >31 days (up to 96 days) after onset of the disease. Clinical sensitivity of NP-IgG was 59.3% at 7–9 days, that of NP-Total Ig was 74.5%, that of SP-IgG was 40.7% and that of SP-Total Ig was 37.0%, which indicated that NP-Total Ig was higher sensitivity than the other antibodies in early phase of COVID-19. (Figure 2A). All the four antibody titers increased around 13 days after symptoms onset respectively (Figure 2B-E).

**Figure 2.**
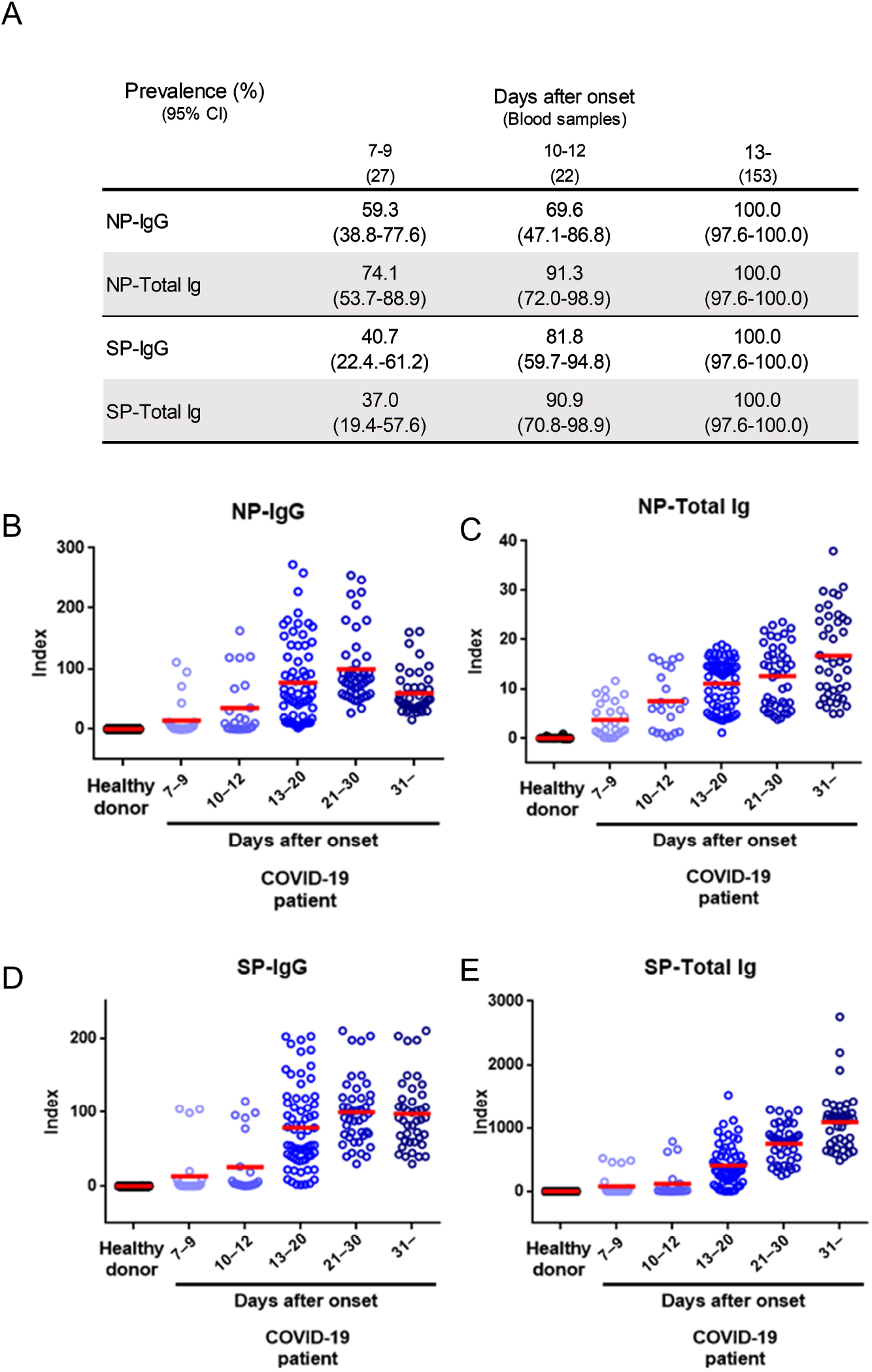
Performance characteristics of NP-IgG, NP-Total Ig, SP-IgG and SP-Total Ig tests in the TOSOH AIA-CL system (A) Summary of sensitivity of each antibody test at the indicated number of days after symptom onset in 202 serum samples from 42 COVID-19 patients. (B) Dot plots of index values of NP-IgG, (C) NP-Total Ig, (D) SP-IgG and (E) SP-Total Ig at the indicated number of days after symptom onset. NP: nucleocapsid protein, SP: spike protein, AIA-CL: automated chemiluminescent enzyme immunoassay system

Furthermore, chronological observation of antibody titers of serial samples collected from nine representative patients are shown in Figure 3. NP-IgG titer was slightly decreased at more than 30 days after symptoms onset (Figure 3A), but NP-Total Ig, SP-IgG and SP-Total Ig titers were increased continuously for up to 21-30 days after symptoms onset and tended to be sustained (Figure 3B-D). There was no obvious difference of the transition of each antibody titer among moderate, severe and critical cases (Figure 3A-D).

**Figure 3.**
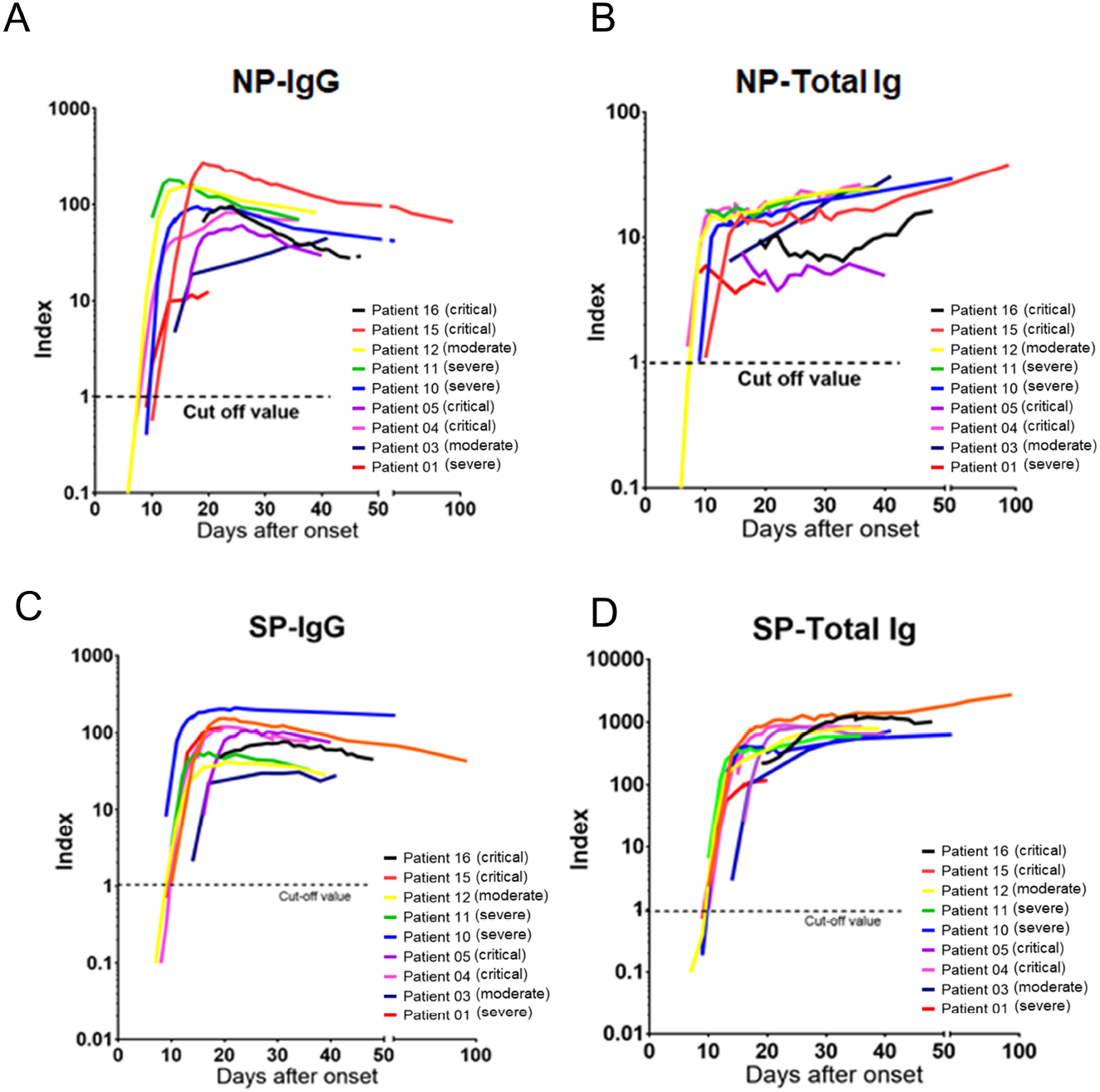
Chronological analysis of antibody titers for representative COVID-19 cases (A) The time course of NP-IgG, (B) NP-Total Ig, (C) SP-IgG and (D) SP-Total Ig index values from symptom onset is shown for nine representative individuals. Each individual is represented by a different color. A 1.0 index cutoff value for positivity is depicted by a horizontal dashed line. The severity is based on NIH classification. NIH: National Institutes of Health

### Sensitivity and specificity of NP-IgG, NP-Total Ig, SP-IgG and NP-Total Ig assays

We compared antibody titers of samples from 1,000 healthy donors and 153 samples from COVID-19 patients collected 13 days after symptoms onset and analyzed using the NP-IgG, NP-Total Ig, SP-IgG and SP-Total Ig reagents (Figure 4A-D). Subsequently, we determined the sensitivity and specificity of our antibody assays by ROC analysis. Each of the four antibodies precisely identified all the true negatives and true positives thereby achieving 100% sensitivity and specificity (Figure 4E-H). Given its excellent performance, our newly developed assay may be suitable for large-scale antibody detection.

**Figure 4.**
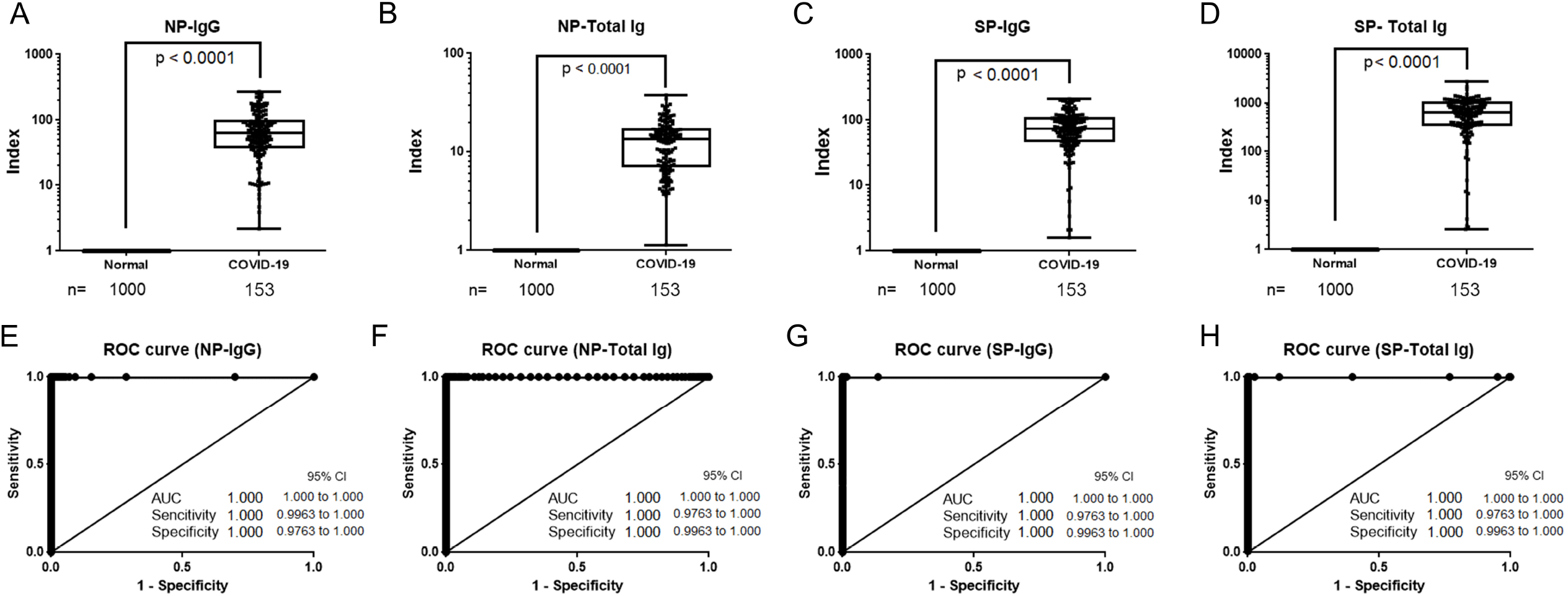
Sensitivity and specificity of TOSOH NP-IgG, NP-Total Ig, SP-IgG and SP-Total Ig AIA-CL reagents (A) Box plots for NP-IgG, (B) NP-Total Ig, (C) SP-IgG and (D) SP-Total IG tests. A total of 153 samples (>13 days after symptoms onset) from 30 patients hospitalized in YCUH and KCRC, and 1,000 healthy donor samples were used. (E) ROC curves of NP-IgG, (F) NP-Total Ig, (G) SP-IgG and (H) SP-Total Ig tests are shown. NP: nucleocapsid protein, SP: spike protein, ROC: receiver operating characteristic, AIA-CL: automated chemiluminescent enzyme immunoassay system, YCUH: Yokohama City University Hospital, KCRC: Kanagawa Cardiovascular and Respiratory Center.

### Correlation with other commercially available kits

We next wanted to compare the performance our test against the commercially successful chemiluminescent Immunoassay platforms Roche-Total Ig and Abbott IgG. As these platforms utilize only the NP, we examined the correlations between the data acquired using the NP-IgG and NP-Total Ig tests from this study and the data acquired using the above mentioned commercial kits. For this purpose, we used 44 COVID-19 antibody–positive samples. Results from NP-IgG were strongly correlated with the results from Roche-Total Ig (Pearson’s correlation coefficient, r=0.97) and Abbott IgG (r=0.962), as were the results from NP-Total Ig (r=0.989 and 0.875, respectively) (Table). Because our antibody tests can measure both NP-IgG and NP-Total Ig with high reliability and additionally SP-IgG and SP-Total Ig, they have a practical scope for COVID-19 serological analysis.

**Table.** Correlations with positivity rates of other commercially available kits for COVID-19 antibody using Pearson’s correlation coefficient NP: nucleocapsid protein, SP: spike protein

| Kit | n | TOSOH<br>NP-IgG | TOSOH<br>NP-Total Ig | Roche<br>NP-Total Ig | Abbott<br>NP-IgG |
| --- | --- | --- | --- | --- | --- |
| TOSOH NP-IgG | 44 |  | 0.889* | 0.970* | 0.962* |
| TOSOH NP-Total Ig | 44 |  |  | 0.989* | 0.875* |
| Roche NP-Total Ig | 11 |  |  |  | 0.975* |
| Abbott NP-IgG | 29 |  |  |  |  |

\* p<0.001
NP: nucleocapsid protein, SP: spike protein

## Discussion

COVID-19 being a global public health problem, surveillance to estimate infection rates based on percentage of antibody positivity and chronological analysis of antibody titers are actively being conducted (18). This is done either as a qualitative assessment using a rapid diagnostic kit or as a quantitative evaluation by antibody titer measurement using ELISA. In particular, the large-scale evaluation of seroprevalence and herd immunity against SARS-CoV-2 requires a high-throughput and quantitative screening as it is more rapid and accurate measurement of viral antibody levels. In this study, we established a combination of NP-IgG, NP-Total Ig, SP-IgG and SP-Total Ig tests for use with the TOSOH AIA-CL system. The sensitivity and specificity of each of the four antibody assay individually was 100% in COVID-19 patient samples collected 13 days after symptoms onset. Moreover, the NP-IgG and NP-Total Ig assays correlated strongly with Abbott and Roche assays for COVID-19 serodiagnosis (19, 20, 21). Considering its excellent performance, this assay could play important roles in generating authentic data during public surveillance of COVID-19.

All the four types of antibody titers began to rise 7–9 days after symptoms onset and were positive in 100% of patients after 13 days, which is consistent with previous reports (4, 22, 23). In addition, antibodies against NP had higher sensitivity than antibodies against SP in the early phase of COVID-19 infection and NP-Total Ig showed the highest sensitivity among the four antibodies. It has been reported that the positivity of NP antibodies is higher than SP antibodies in acute phase of SARS-CoV-2 infection (24, 25). As in common viral infections, COVID-19 infection also has polyclonal elevation of Immunoglobulin such as IgM and IgA, not only IgG. Although many antibody kits can measure only IgG or/and IgM, it has been reported that measuring IgA in addition to IgG or IgM increases the sensitivity of the assay (8). Since our assay measures total-Ig in addition to IgG, it can possibly be more precise than assays that only detect a particular Ig type.

Antibodies against spike antigen, especially RBD, have the ability to prevent the entry of the virus into the host cells (26, 27). According to previous reports, while NP-IgG declined slowly over the course of approximately half a year and turned negative in some cases, SP-IgG declined slowly but sustained its antibody titer and rarely turned negative (28). In this study, all the four antibodies were positive in all cases after 13 days from symptoms onset, and NP-Total Ig, SP-IgG, and SP-Total Ig have been sustained during the period of observation. However, NP-IgG titers tended to decrease after 30 days from symptoms onset (Figure 4) similar to the trend noted by Long et al., who observed the decline of IgG antibodies in convalescence (29). Multiple antibody measurements, especially antibodies against SP, are likely to be useful for assessment of long-term changes of antibody titers and providing important insights into immune memory.

It should be noted that there were some limitations of this study. First, most cases in this study were of high severity. It is known that COVID-19 antibody titers tend to be lower in less severe cases (29, 30). Although there was no obvious difference in the antibody titers between moderate, severe and critical cases in this study, it was insufficient to evaluate the different in antibody titers due to the severity of COVID-19 because the number of samples in moderate cases were smaller than that of severe or critical cases. Therefore, there is the possibility that the cutoff value set in this assay may not be appropriate for detecting the antibody titers in lower severity cases, such as mild or asymptomatic patients. Second, the newly developed reagents can only measure IgG and total Ig, but not other immunoglobulin levels, such as IgM, IgA. Also, this study showed that NP-Total Ig was sustained but NP-IgG decreased after 30 days since symptoms onset. The reason for this reduction in IgG titers is unclear and the other immunoglobulin which elevated to sustain the total Ig levels also remains to be identified with further investigations.

In conclusion, the automated TOSOH AIA-CL assay that can simultaneously quantify anti-SARS-CoV-2 NP-IgG, NP-Total Ig, SP-IgG and SP-Total Ig which was developed in this study, performed to be a highly accurate and specific serodiagnostic test for COVID-19. This test could play an important role in accurate detection of COVID-19 patients, evaluation of chronological changes in multiple antibody titers and in large-scale sero-surveillance studies.

## Data Availability

The data used in this study are available upon request as they contain potentially identifying or sensitive patient information. This policy follows the restrictions imposed by Yokohama City University Hospital and Kanagawa Cardiovascular and Respiratory Center institutional review board that approved the study. Electronic health record data cannot be shared publicly because they consists of personal information from which it is difficult to guarantee de-identification. As a result, there is a possibility of deductive disclosure of participants and therefore full data access through a public repository is not permitted by the institutions that provided us the data. The data and associated documentation from each collaborating institution can only be made available under a new data sharing agreement with which includes: 1) commitment to using the data only for research purposes and not to identify any individual participant; 2) a commitment to securing the data using appropriate measures, and 3) a commitment to destroy or return the data after analyses are complete. Requests can be made directly at aryo @yokohama-cu.ac.jp

## Acknowledgments

We thank Chizu Suzuki, Natsumi Takaira, Kenji Yoshihara, and Kazuo Horikawa for their technical assistance.

## Contribution

S.K., N.O. analyzed the data, verified the analytical methods and wrote the manuscript, S.S.J, Kei Miyakawa. analyzed the data and wrote the manuscript, Y.Y., T.M., A.G., T.K., T.Y., analyzed the data, Kota Murohashi, E.H., E.Y., T.O. contributed reagents, A.R. directed the research, analyzed the data and wrote the manuscript.

---

**Figure S1.** Determination of cutoff values of NP-IgG, NP-Total Ig, SP-IgG and SP-Total Ig tests in the TOSOH AIA-CL system

(A) Comparison of NP-IgG, (B) NP-Total Ig, (C) SP-IgG and (D) SP-Total Ig titers between COVID-19 patients and non-infectious healthy donors in the TOSOH AIA-CL system. 17 serum samples derived from COVID-19 patients and 600 samples from non-infectious healthy donors were used for analysis. Statistical analysis was performed by Mann–Whitney Rank-Sum test, and p-values are presented above the box plots. (E) ROC curves for NP-IgG, (F) NP-Total Ig, (G) SP-IgG and (H) SP-Total Ig tests as measured in (A) to (D). Cutoff values at 1.0 index were 3,889 counts of intensity for NP-IgG and 1,705 counts for NP-Total Ig, 6,700 counts for SP-IgG, 1,000 counts for SP-Total Ig.

NP: nucleocapsid protein, SP: spike protein, AIA-CL: automated chemiluminescent enzyme immunoassay system, ROC: receiver operating characteristic

**Table S1.** Characteristics of healthy donors and COVID-19 patients

|  | Healthy donors | COVID-19 patients |  |
| --- | --- | --- | --- |
|  |  | YCUH | KCRC |
|  | n (Blood samples) | n (Blood samples) | n (Blood samples) |
| Gender |  |  |  |
| Male | 323 (323) | 16 (161) | 19 (19) |
| Female | 677 (677) | 2 (15) | 7 (7) |
| Mean age [SD] | 33.1 [7.6] | 61.3 [16.5] | 64.5 [17.2] |
| Severity |  |  |  |
| Asymptomatic |  | 0 | 0 |
| Mild |  | 1 (1) | 1 (1) |
| Moderate |  | 3 (20) | 14 (14) |
| Severe |  | 8 (65) | 8 (8) |
| Critical |  | 6 (90) | 1 (1) |
| Others |  | 0 | 2 (2) |

YCUH: Yokohama City University Hospital, KCRC: Kanagawa Cardiovascular and Respiratory Center.

