## Supplementary figures and images for "Development of an automated chemiluminescence assay system for quantitative measurement of multiple anti-SARS-CoV-2 antibodies"

### Supplemental Figure 1

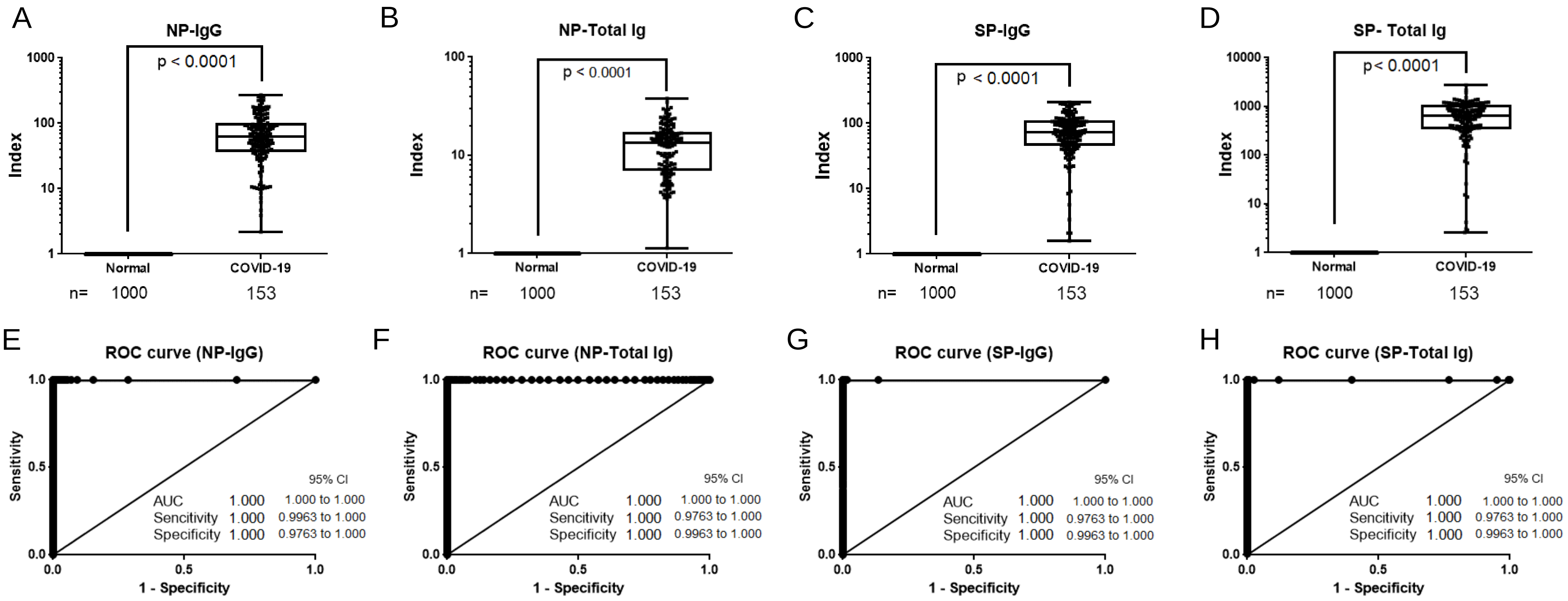
